# The Alberta Network for Community Health Outreach and Rural Mental Health (ANCHOR-MH): A pilot study of a collaborative educational initiative to improve psychiatric outcomes in primary care

**DOI:** 10.64898/2026.01.08.25342158

**Authors:** Jeremy Weleff, Evan J. Kyzar, Hannah Pazderka, Mayada Akil, Andrew Baxter, Alberto L. Choy, Joseph J. Cooper, Adriane dela Cruz, Jane L. Eisen, Brady John Heward, Sheny Khera, Christina Korownyk, Mobolaji A. Lawal, Eden McCaffrey, Chantal Moreau, Daniel Moreno De Luca, Emma Samelson-Jones, Adegboyega Sapara, Gaurav Sharma, Yifeng Wei, Avery Wynick, Bernice N. Yau, Yanbo Zhang, David A. Ross

**Author notes:** Corresponding author Abstract word count: 250. Co-first authors (Jeremy Weleff, Evan J. Kyzar).

## Abstract

**Background:** Approximately 1 in 5 Canadians experience a mental health illness in any given year. While most individuals can be successfully treated within a primary care setting, a subset of individuals present with a severity and complexity requiring specialist care. Unfortunately, a shortage of psychiatrists (especially in rural regions) can result in wait times of months to years.

**Methods:** We designed the Alberta Network for Community Health Outreach and Rural Mental Health. ANCHOR-MH is a 12-week program that includes a unique educational intervention, collaborative case conferencing, and a community of practice between family medicine (FM) physicians and psychiatrists. We enrolled two pilot cohorts of n=20 FM physicians each and measured participants’ confidence and comfort in diagnosing, managing, and treating psychiatric conditions. We also conducted qualitative analyses of their experience.

**Results:** Data from participants that completed both the pre- and post-program survey (n=34) showed increased confidence in screening for, diagnosing, and managing psychiatric issues, as well as increased comfort discussing mental health concerns with patients and families and reduced stigma towards certain psychiatric conditions. Qualitative thematic analysis (n=39) reflected this increased confidence, revealed an increased sense of connectedness to the mental healthcare landscape, and highlighted specific examples of practice changes. Participants broadly agreed that the program improved their ability to provide mental healthcare and would improve psychiatric outcomes within their practice.

**Interpretation:** ANCHOR-MH improved FM physicians’ confidence and ability to deliver mental healthcare in their primary care settings. Increasing the reach of this program may improve mental healthcare in underserved communities.

## Introduction

Approximately 1 in 5 Canadians experience a mental health problem in any given year and, by age 40, approximately 50% of individuals will have experienced a mental illness (1,2). Beyond the human distress caused by these conditions, the economic burden of mental illness in Canada is estimated at $51 billion per year (2). Most Canadians rely on their primary care providers to address their mental health needs. While this may be effective for many patients, there is a subset of individuals who present with a severity and complexity that requires specialist care. Unfortunately, a shortage in the number of psychiatrists – as well as an imbalance in their geographical distribution, particularly in rural and northern Canada (3–5) – can result in wait times of months to years (6,7). In the meantime, primary care teams must manage these cases in settings not designed for specialized psychiatric care, creating a mismatch that can lead to provider frustration or moral injury and potentially suboptimal patient outcomes.

Emerging evidence suggests that educational initiatives (such as Canadian Research and Education for the Advancement of Child Health (CanREACH) and Extension for Community Healthcare Outcomes (ECHO) (8,9)) and innovative ways of delivering psychiatric care (such as electronic consult services (10) and collaborative care models (11)) may help address this tension. While each of these approaches has their strengths, there may be room to optimize pedagogy and to create synergy from the best practices of each. To this end, we designed ANCHOR-MH (Alberta Network for Community Health Outreach and Rural Mental Health), a novel educational program with the goal of enhancing Family Medicine (FM) physicians’ confidence and skill at treating psychiatric illnesses, improving collaboration between FM physicians and psychiatrists, and establishing an enduring community of practice.

## Methods

### Intervention design and setting

ANCHOR-MH is a 12-week program designed to offer family physicians the opportunity to deepen their expertise and leadership in mental healthcare and to enhance long-term collaboration between FM physicians and psychiatrists. The curriculum includes two hours of online synchronous activity each week^1^, conducted via Zoom, with asynchronous engagement via a private Discord community. Each cohort operates in a tri-partite model that balances directed learning with real-world clinical application (Figure 1):

1. *Structured experiential learning*. The first hour of each week offers clinically focused, high-yield content relating to a particular psychiatric diagnosis using a framework that leverages core principles of adult learning (including being case-based, experiential, and with rigorous formative assessment; see Figure 1) (12). The sessions cover key content (e.g. treatment guidelines and core pathophysiology across a range of diagnoses; Supplemental Table 1) while also highlighting critical elements of the overall approach to the psychiatric patient (e.g., navigating diagnostic complexity, countertransference).
2. *Case conferencing.* A major challenge of any medical educational initiative is to translate classroom learning into clinical practice. To this end, the second hour of each week is built around collaborative case work, with groups of five family physicians consulting with two (or more) psychiatrists regarding current patients they treat for mental health concerns. This structure offers personalized instruction and supports complex decision-making.
3. *Community of practice.* To further assist knowledge translation and bridge structural gaps, an asynchronous platform was used to share resources and through which participants could discuss ongoing questions with peers and consultants. This structure also enables ongoing support, case conferencing, and peer-to-peer mentoring for FM physicians beyond the initial 3-month experience.

**Figure 1.**
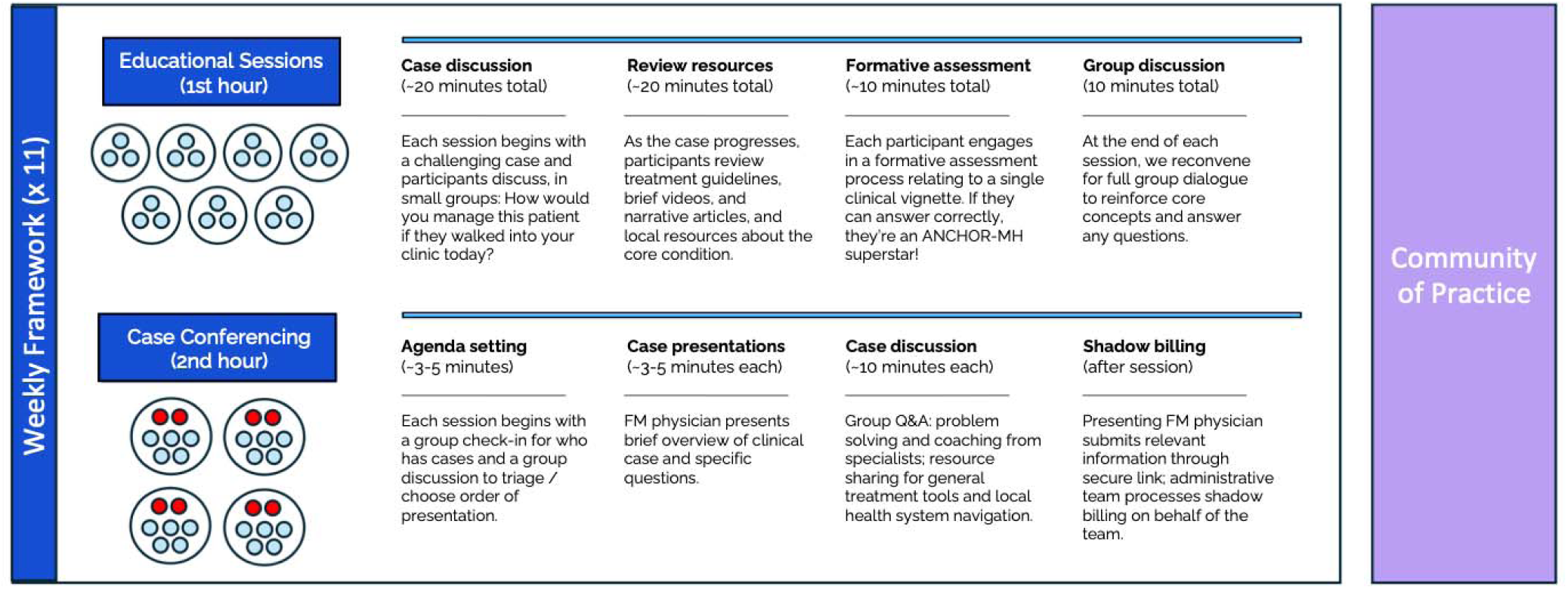
Schematic of ANCHOR-MH program. The program consists of a tri-partite structure: 1) one-hour, small group (n = 3-4), experiential learning exercise with FM physicians; 2) one-hour, small group case-conferencing with FM physicians (n=5) and psychiatrists (n=2-3); 3) community of practice including asynchronous discussion (via Discord) and ongoing case-conference opportunities.

### Recruitment

For our initial cohort (Cohort 1), a call for applications was sent through the Department of Family Medicine clinical faculty listserv. n=42 FM physicians applied for the program and n=20 were accepted based on review of their CV and statement of interest; all n=20 completed the program from January to March of 2025. For Cohort 2, a call for applications was sent through the same listserv and shared by previous participants through their social networks. n=109 FM physicians applied and n=20 were accepted based on review of materials; one additional participant was accepted as a liaison to the Department of Family Medicine. n=20 individuals completed the program from July to September of 2025 (one individual could not participate because of a family conflict).

### Quantitative Data Collection & Analysis

Participants were asked to complete a pre- and post-survey that included questions about their FM practice setting, past training in psychiatry, and an assessment of the burden of psychiatric cases in their practice. We assessed 5 co-primary outcomes, including participants’ ratings of the following items: 1) “I feel confident in my ability to screen for and diagnose common psychiatric conditions in my practice”; 2) “I feel confident in my ability to manage common psychiatric conditions in my practice”; and 3) “I feel comfortable discussing mental health concerns with my patients and their families” on a 6-point Likert scale (1=Strongly disagree; 2=Disagree; 3=Not sure but probably disagree; 4=Not sure but probably agree; 5=Agree; 6=Strongly agree). The other 2 co-primary outcomes were attitudes towards patients with 4) borderline personality disorder, a condition known to have significant stigma (13), and 5) hypertension, as a medical control. Attitudes were measured using the Medical Condition Regard Scale (MCRS) (14) in both the pre- and post-surveys. Data were analyzed with paired, two-sided Student’s *t*-tests corrected for multiple comparisons (critical α=0.01). Wilcoxon signed-rank tests were used for sensitivity analysis, yielding nearly identical statistical results. Incomplete or incorrectly filled surveys were excluded from the evaluation (n=1 from pre-survey (Cohort 1); n=5 from post-survey (n=4 from Cohort 1)). Data were combined across cohorts for the main analyses.

In the post-survey, participants used a 6-point Likert scale to rate whether the program: 1) was a good use of their time; 2) improved their ability to care for psychiatric patients; 3) improved their knowledge of local psychiatric resources; 4) increased their confidence in their patients receiving better care; and 5) whether they would refer colleagues to the program. Data were analyzed in Python v3.10.12 utilizing NumPy (15) and SciPy (16).

### Qualitative Interviews & Analysis

The post-survey for Cohort 2 included spaces for free-text responses, including: “Have you changed your approach to patient care in any way? (if yes, how?)”, “Do you have any specific feedback on the education sessions we ran in the first hour of each week?”, and “Do you have any specific feedback on the case conferencing component?”.

Qualitative interviews were also held virtually prior to beginning the experience and then again within 1 month of program completion, with the goal of understanding individuals’ goals for the experience (pre-interview) and then general feedback on the structure and efficacy of the program (post-interview). Interviews were requested with all participants and n=39 took part. Discussions followed a semi-structured interview guide (Supplemental Material). Thematic analysis (17) was conducted by a single researcher separately for each cohort, with confirmatory analysis of the themes by 3 additional researchers. The data sets were coded independently of one another, to allow for the possibility of new or emergent themes arising in the different cohorts.

### Funding

Funds for an initial pilot were obtained from the existing Academic Medicine and Health Services Program (AMHSP) budget from the Departments of Family Medicine and Psychiatry at the University of Alberta. The AMHSP is an Alternative Remuneration Plan (ARP) designed to support academic medicine in the province (including research, education, and clinical innovation). Both FM physicians and psychiatrists were compensated for their time at a standard clinical rate; the protected academic time was offset by “shadow billing” during the case conferencing.

### Ethics

The project was designed with the intent of improving provincial health service delivery by evaluating the efficacy of an educational intervention, and, as such, we received a letter of exemption from the Health Research Ethics Board (HREB) at the University of Alberta.

## Results

### Participant information

Baseline information from n=39 FM physicians who took part in ANCHOR-MH and completed the pre-survey is shown in Table 1. Physicians had a range of practice experience, and most (76.9%) worked in an urban setting. The vast majority (94.9%) reported receiving between 2-8 weeks of supervision under a psychiatrist during FM residency. The estimated number of daily visits involving psychiatric and/or addiction concerns varied but was generally high (with 38.5% of FM providers stating that >41% of their daily case load involved these concerns), and a similar pattern was noted for the percentage of participants’ patient panels with unmet psychiatric needs. Participants estimated the wait time for psychiatric referrals in their practice was substantial, with nearly half of participants (43.6%) estimating greater than 6 months.

**Table 1.** Baseline participant information.

| **Question** | **Cohort 1 (n=19)**<br>**n (% of total)** | **Cohort 2 (n=20)**<br>**n (% of total)** | **Total (n=39)**<br>**n (% of total)** |
| --- | --- | --- | --- |
| *Years in practice* |  |  |  |
| < 5 years | 6 (31.6) | 6 (30) | 12 (30.8) |
| 5-10 years | 5 (26.3) | 5 (25) | 10 (25.6) |
| 11-15 years | 0 | 7 (35) | 7 (17.9) |
| 16-20 years | 3 (15.8) | 1 (5) | 4 (10.3) |
| > 20 years | 5 (26.3) | 1 (5) | 6 (15.4) |
| *Practice setting* |  |  |  |
| Urban | 17 (89.5) | 13 (65) | 30 (76.9) |
| Suburban | 2 (10.5) | 3 (15) | 5 (12.8) |
| Rural | 0 | 3 (15) | 3 (7.7) |
| Remote | 0 | 1 (5) | 1 (2.6) |
| *Do you supervise trainees?* |  |  |  |
| Yes | 16 (84.2) | 18 (90) | 34 (87.2) |
| No | 3 (15.8) | 2 (10) | 5 (12.8) |
| *Amount of psychiatry-specific training in FM residency* |  |  |  |
| < 2 weeks | 0 | 0 | 0 |
| 2-4 weeks | 8 (42.1) | 16 (80) | 24 (61.5) |
| 5-8 weeks | 10 (52.6) | 3 (15) | 13 (33.3) |
| 9-12 weeks | 0 | 0 | 0 |
| > 3 months | 1 (5.3) | 1 (5) | 2 (5.1) |
| *Percentage of daily patient visits involving psychiatric or addiction concerns* |  |  |  |
| 0-20% | 1 (5.3) | 7 (35) | 8 (20.5) |
| 21-40% | 7 (36.8) | 9 (45) | 16 (41) |
| 41-60% | 4 (21.1) | 1 (5) | 5 (12.8) |
| 61-80% | 3 (15.8) | 3 (15) | 6 (15.4) |
| > 80% | 4 (21.1) | 0 | 4 (10.3) |
| *Percentage of patient panel having unmet psychiatric or addiction needs* |  |  |  |
| 0-20% | 1 (5.3) | 4 (20) | 5 (12.8) |
| 21-40% | 5 (26.3) | 10 (50) | 15 (38.5) |
| 41-60% | 6 (31.6) | 2 (10) | 8 (20.5) |
| 61-80% | 2 (10.5) | 3 (15) | 5 (12.8) |
| > 80% | 5 (26.3) | 1 (5) | 6 (15.4) |
| *Estimated wait time for psychiatrist after referral* |  |  |  |
| < 1 month | 1 (5.3) | 2 (10) | 3 (7.7) |
| 1-3 months | 3 (15.8) | 2 (10) | 5 (12.8) |
| 3-6 months | 6 (31.6) | 8 (40) | 14 (35.9) |
| 6-12 months | 7 (36.8) | 4 (20) | 11 (28.2) |
| >12 months | 2 (10.5) | 4 (20) | 6 (15.4) |
| *Assessment of current access to psychiatric care for patient panel* |  |  |  |
| Very poor | 0 | 3 (15) | 3 (7.7) |
| Poor | 10 (52.6) | 8 (40) | 18 (46.2) |
| Not sure but probably poor | 5 (26.3) | 4 (20) | 9 (23.1) |
| Not sure but probably good | 1 (5.3) | 2 (10) | 3 (7.7) |
| Good | 1 (5.3) | 2 (10) | 3 (7.7) |
| Very good | 2 (10.5) | 1 (5) | 3 (7.7) |

### Impact of ANCHOR-MH on co-primary outcomes

Participants’ confidence in their ability to screen for, diagnose, and treat common psychiatric conditions, as well as their comfort discussing mental health concerns with patients and families, was significantly increased in the post-survey (Table 2). Participation in the program improved attitudes towards individuals with borderline personality disorder using the MCRS; MCRS scores for hypertension did not meet multiple comparisons correction criteria for significance. All participants who completed the post-survey (n=35) felt the program was a good use of their time and that the program improved their ability to care for their patients’ psychiatric needs (35/35 agree or strongly agree, 100%; Figure 2). Most felt that their patients will receive better care because of this program (33/35 agree or strongly agree, 94.3%; 2/35 not sure but probably agree, 5.7%) and that the program increased their knowledge of local psychiatric resources (28/35 agree or strongly agree, 80%; 6/35 not sure but probably agree, 17.1%; 1/35 not sure but probably disagree, 2.9%). Participants felt positively about referring colleagues to the program (33/35 agree or strongly agree, 94.3%; 2/35 not sure but probably agree, 5.7%).

**Figure 2.**
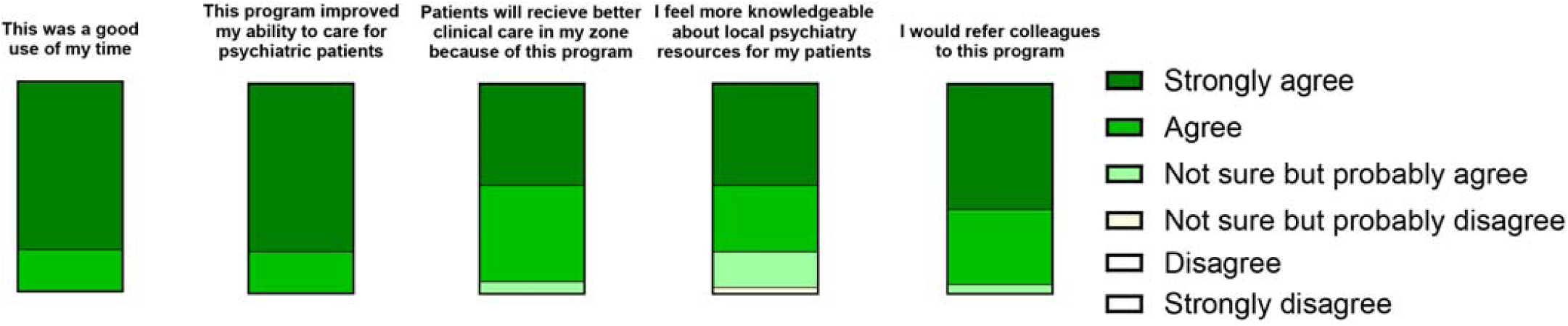
Post-survey program assessment. Participants were asked the questions shown in the figure on a 6-point Likert scale (Strongly disagree; Disagree; Not sure but probably disagree; Not sure but probably agree; Agree; Strongly agree). Of note, no participants responded “Disagree” or “Strongly disagree” for any question.

**Table 2.** Comparison of confidence and comfort managing psychiatric conditions and medical condition regard scale (MCRS) scores pre- and post-intervention. Note that the maximum total value possible for the confidence/comfort measures is equal to 6, while the total possible MCRS range is 11-66. *Asterisk indicates that measure meets critical *p*-value (*p*<0.01) after correction for multiple comparisons. n=34.

| Co-primary outcome | Pre-survey mean ± SD | Post-survey mean ± SD | $t$ statistic | $p$-value |
| --- | --- | --- | --- | --- |
| *Confidence/comfort measures* |  |  |  |  |
| I feel confident in my ability to screen for and diagnose common psychiatric conditions in my practice | 4.9 ± 0.6 | 5.5 ± 0.5 | -5.52 | 0.000003* |
| I feel confident in my ability to manage common psychiatric conditions in my practice | 4.7 ± 0.7 | 5.4 ± 0.5 | -5.14 | 0.00001* |
| I feel comfortable discussing mental health concerns with my patients and their families | 4.9 ± 0.7 | 5.5 ± 0.5 | -4.89 | 0.00003* |
| *Attitudes towards specific conditions (MCRS)* |  |  |  |  |
| Borderline Personality Disorder | 44.1 ± 8.0 | 47.6 ± 6.1 | -3.43 | 0.002* |
| Hypertension | 49.6 ± 7.2 | 51.9 ± 5.5 | -2.38 | 0.023 |

### Qualitative themes

Of the participants who completed the program, n=39 underwent a qualitative interview to assess the efficacy of the program and identify opportunities for improvement. Key themes from thematic analysis included: increased confidence in managing psychiatric concerns, appreciation of the quality of the resources (particularly those engaging with the neurobiology of psychiatric disorders), and an increased sense of connectedness to colleagues and to the mental healthcare landscape. Participants appreciated the case conferencing and highlighted strategies for further optimizing its structure. A list of identified themes with representative quotes are presented in Table 3 (additional responses in Supplementary Information).

**Table 3.**
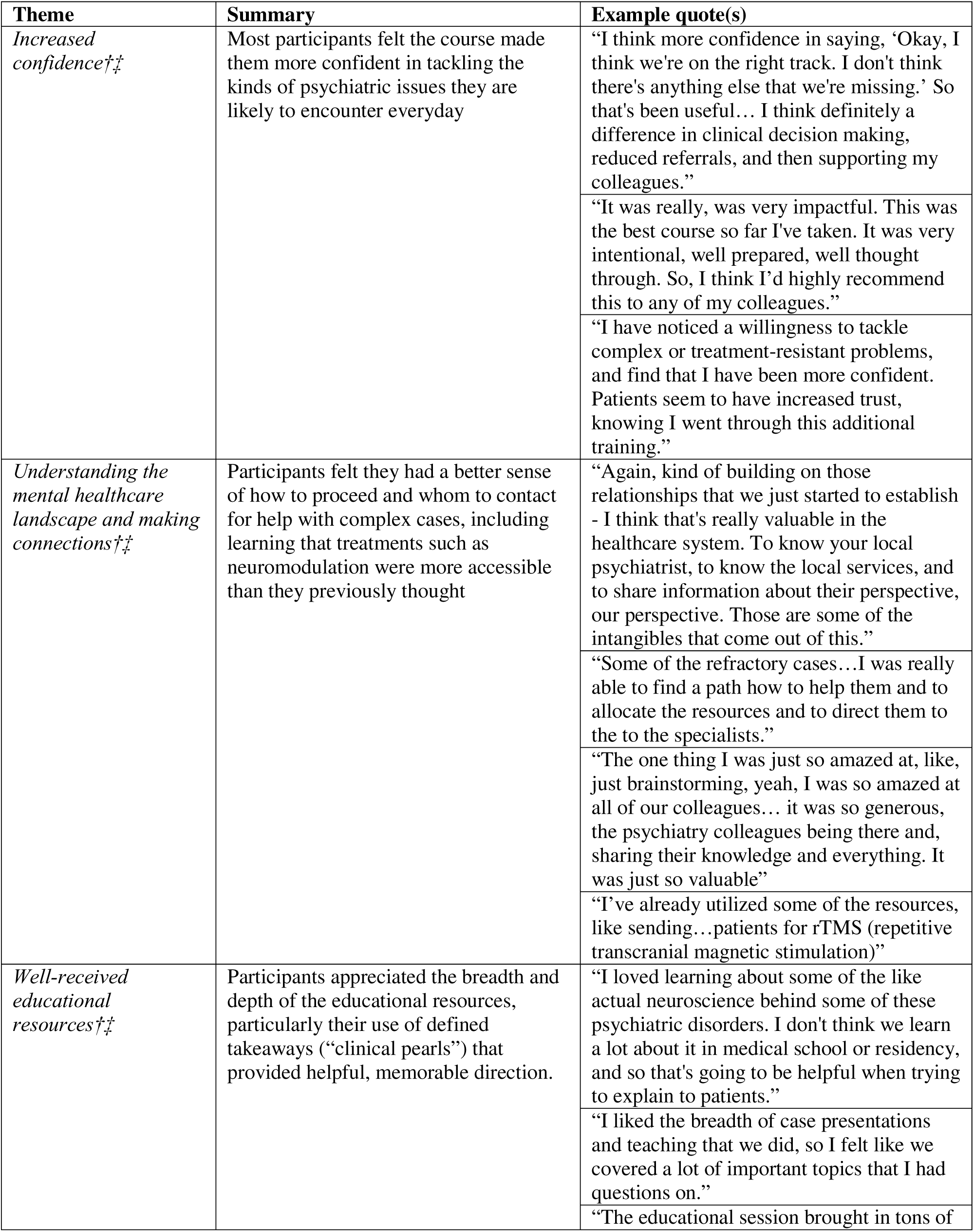
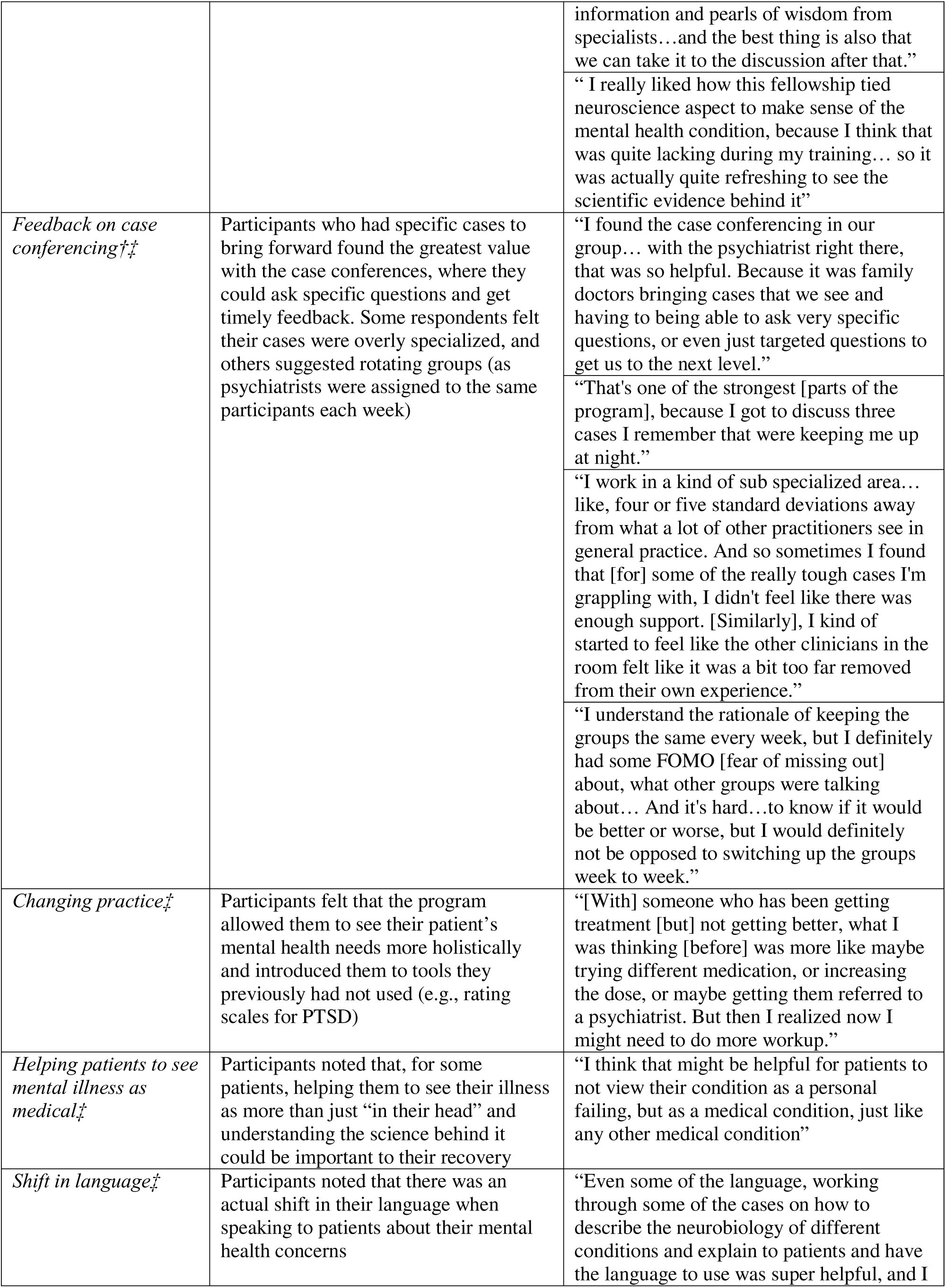
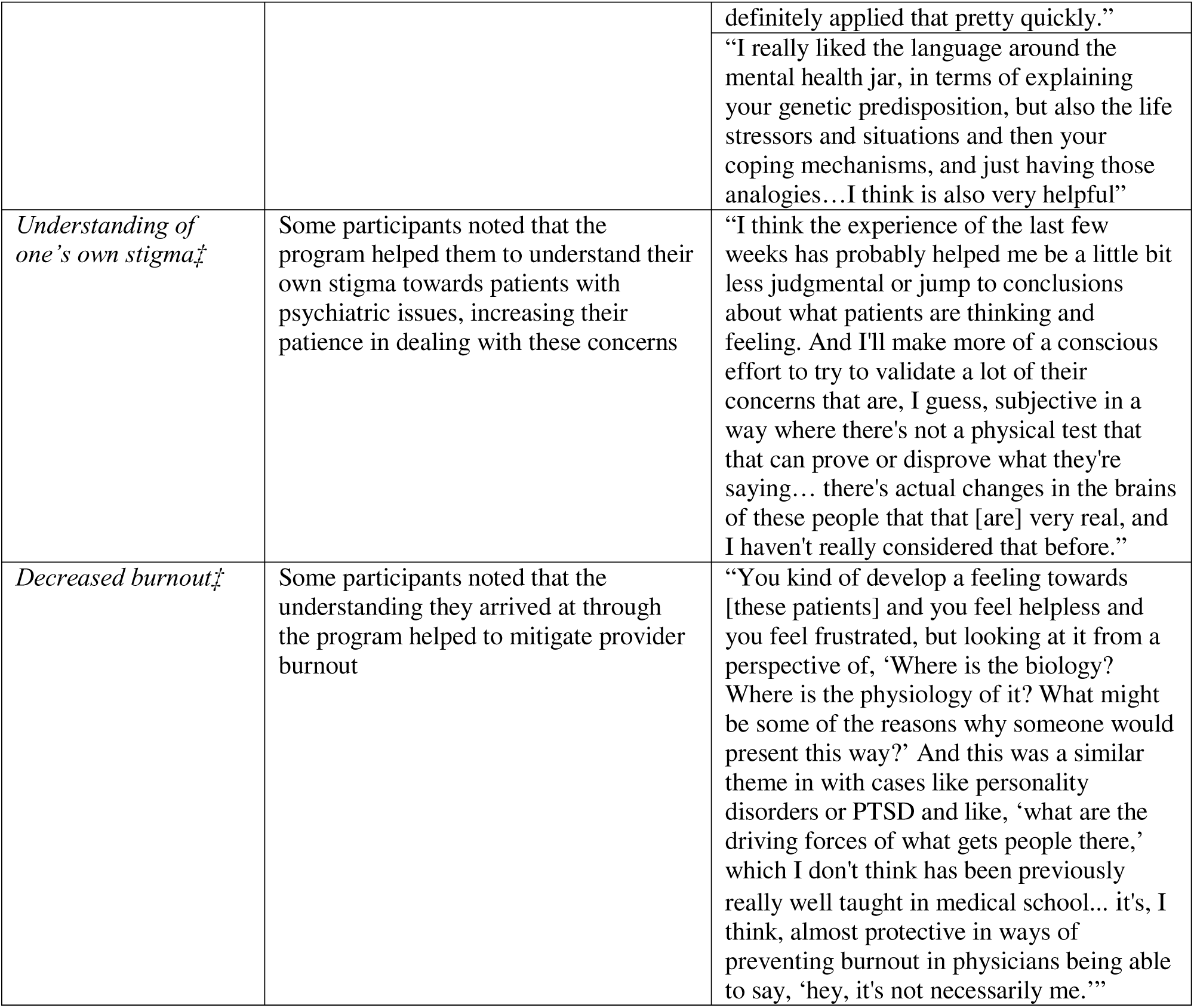
Themes identified from qualitative interviews along with illustrative quotes. †Identified in thematic analysis of Cohort 1; ‡Identified in thematic analysis of Cohort 2. See Supplementary Information for additional representative quotes.

Data from free-text responses in the post-survey with Cohort 2 corroborated these themes. Responses highlighted: how understanding core pathophysiology led to increased empathy towards individuals with psychiatric illness, an appreciation of the educational framework, and specific ways in which some of the FM physicians changed their approach to patient care (sample quotes shown in Table 4; all responses are shown in Supplementary Table 2).

**Table 4.**
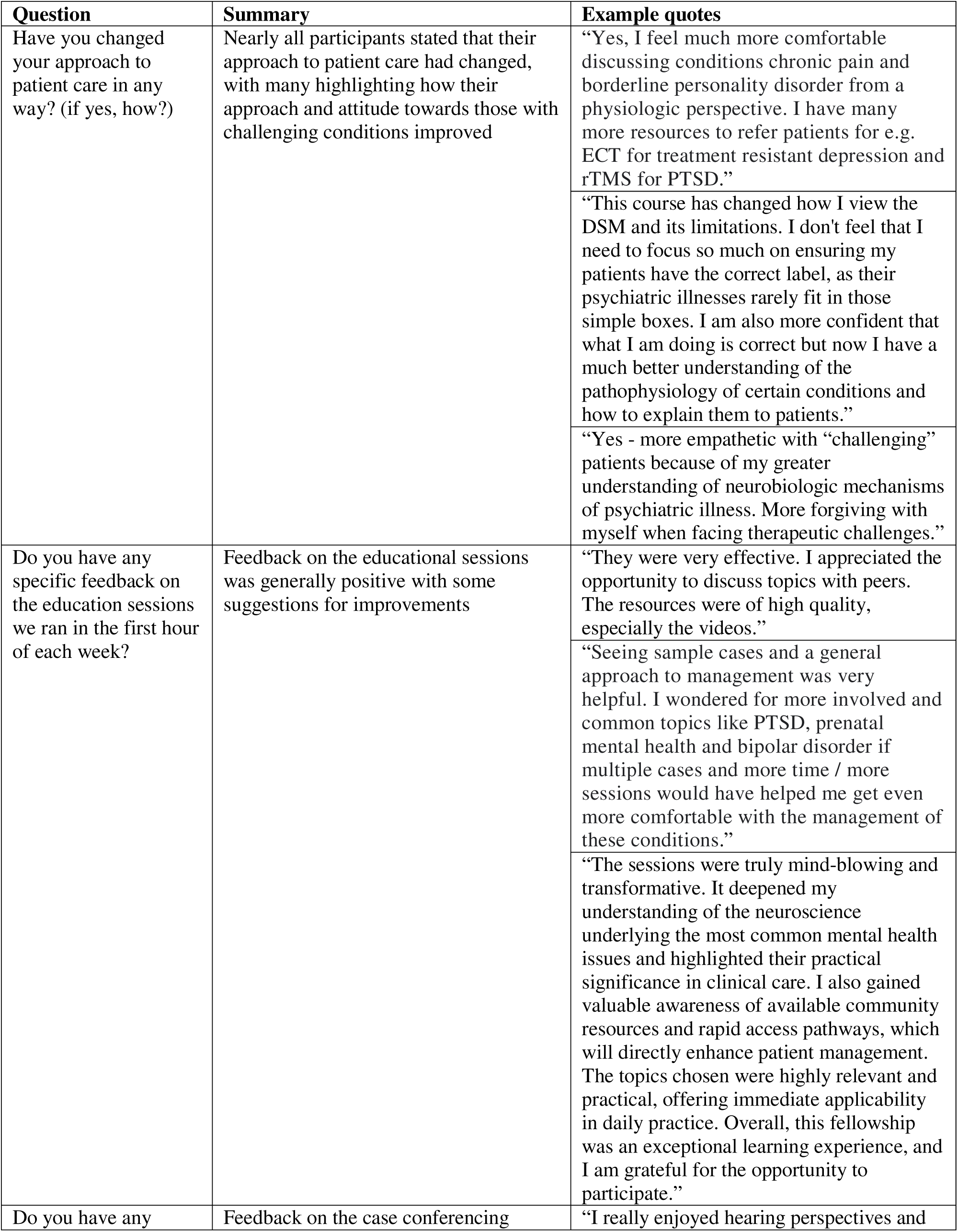
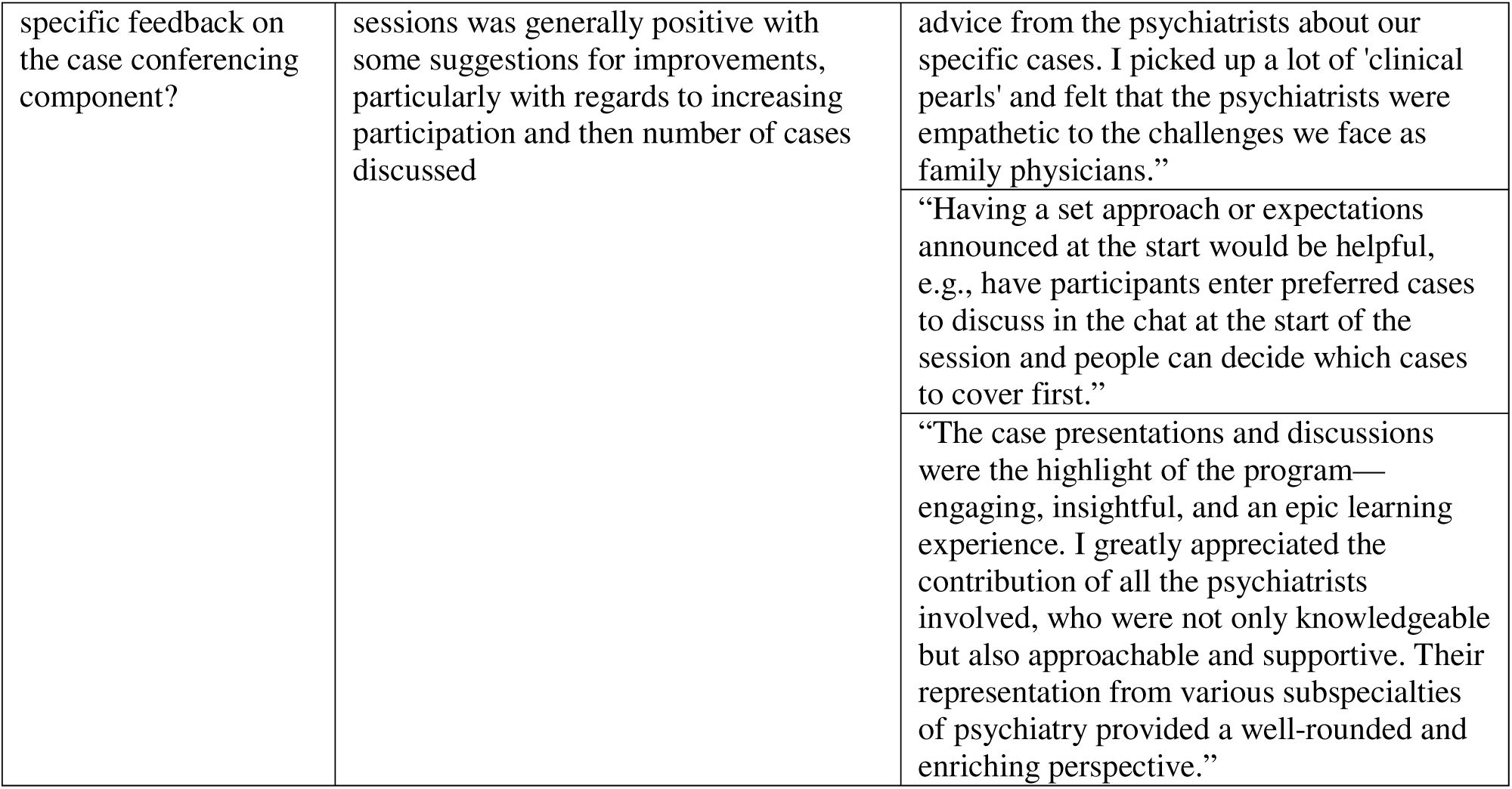
Qualitative summary of data from post-survey questions. See Supplementary Table 2 for additional responses.

## Interpretation

Here, we present a report on a pilot program, ANCHOR-MH, that was designed to address structural gaps in our mental health care system by providing increased training and support for FM physicians and enhancing collaboration between psychiatry and family medicine. Unique aspects included an educational component grounded in principles of adult learning, a case conference designed to facilitate collaboration between FM physicians and psychiatrists, and an asynchronous space designed to build a community of practice.

All participants reported that it would directly improve their ability to care for patients with psychiatric illness. Comparison of pre- and post-surveys showed significant increases in participants’ confidence and comfort in diagnosing, treating, and discussing common psychiatric conditions, and improved attitudes towards individuals with borderline personality disorder.

Qualitative data were striking, including an appreciation for the pedagogy, feeling empowered in their work, and with numerous specific examples of how the program improved their clinical practice. Together, these data suggest that this pilot program was successful and effective.

This program builds on other initiatives, such as CanREACH, ECHO, and others that have been successfully implemented in Canada and elsewhere (8,9,11,18–23). ANCHOR-MH may offer advantages over other programs by virtue of the specific pedagogy, the focus on enhanced local collaboration between FM and psychiatry (with the establishment of an ongoing community of practice), and the innovative financial model (allowing the program to be self-sufficient through a local ARP). This latter point was critical, as fairly compensating physicians for their time avoids exacerbating work pressure in a group that may already feel overburdened (24,25).

Another unique aspect of this program was that many core resources (including a series of clinical commentaries in *Biological Psychiatry*) were designed by the NNCI, a leading team in psychiatric education (26). These pieces distill complex scientific topics and use narrative tools to make them clear, relevant, and accessible to a broad audience. Effective neuroscience education has been shown to reduce stigma toward those with mental illness and/or addiction in psychiatrists (27) and the general public (28). Accordingly, the decrease in MCRS scores towards individuals with borderline personality disorder were especially notable, as were the qualitative data showing that the neuroscientific aspects of the resources were well-received.

The success of this pilot raises the possibility of scaling the program to provide training in regions that have been traditionally underserved and/or have limited access to specialty psychiatric care (4,6,29). While our initial cohort practiced predominantly in an urban setting, the second cohort included ∼25% rural practicing physicians, and upcoming cohorts will focus on recruiting physicians providing care to rural and Indigenous Albertans. Future directions include expanding beyond Alberta (including other rural parts of Canada and internationally). One could also assess whether the program has any direct impact on physicians’ day-to-day clinical practice (e.g. in the resources that they use to help with decision making or in the frequency and nature of referrals they place to specialists) and on the health care system more broadly. Such data would allow for economic and implementation analyses that can influence policy decisions and new models of care (e.g. a collaborative care model through which referrals from FM physicians can be linked to consulting psychiatrists in the program) (11,30–32).

The present study has several limitations. The data are based on a relatively small sample. Moreover, individuals were recruited through connections to an academic department based on their interest in furthering their education and skill in psychiatry; as such, participants may be seen as early adopters, and the data may not generalize to all FM physicians. In considering broader implementation, the program would be somewhat limited by group size. While the case-based learning can easily be run at scale (there is no limit to the number of breakout rooms), the conferencing component is limited by the number of psychiatrists available to participate (and thereby maintain an appropriate ratio). Another potential issue is the program’s reliance on technology, which may limit engagement in rural areas with poor internet access and with providers who are less technologically savvy.

In conclusion, we designed a 12-week program that increased the confidence and ability of FM physicians to provide care for patients with psychiatric and addiction-related concerns while enhancing collaboration between psychiatry and family medicine. Future work will focus on iterating the curriculum (including adding additional resources to the community of practice), enhancing assessment tools, and expanding efforts to address gaps in rural areas and with other underserved communities.

## Conflict of interest disclosures and acknowledgements

EJK is supported by grants from the Leon Levy Foundation and the National Institute on Alcoholism and Alcohol Abuse (L70AA032773). BJH is supported by HRSA-19-108 Rural Communities Opioid Response Program, Rural Centers of Excellence on Substance Use Disorders (RCORP-RCOE). DMDL is supported by the CASA Research Chair. ESJ is supported by Training for Residents Integrating Skills, Treatment Advocacy and Resilience for Mental and Behavioral Health (TRI-STAR-MBH) and the HRSA TA2HP48947-01-00. DAR is supported by the Alberta Health Services Chair in Mental Health Research. The NNCI is funded in part by the Deeda Blair Research Initiative Fund for Disorders of the Brain through support to the Foundation for the National Institutes of Health, with additional funding from the Society of Biological Psychiatry (SOBP) and the American College of Neuropsychopharmacology (ACNP). All authors report no biomedical financial interests or potential conflicts of interest related to the present work.

## Data Availability

All data produced in the present study are available upon reasonable request to the authors.

## Footnotes

1 In the first cohort, the first and last sessions were held in person.

