## Supplemental Information for "The Alberta Network for Community Health Outreach and Rural Mental Health (ANCHOR-MH): A pilot study of a collaborative educational initiative to improve psychiatric outcomes in primary care"

Additional information and quotes from Cohort 1 qualitative interviews……………………….8
Additional information and quotes from Cohort 2 qualitative interviews……………………….12

### Supplementary Table 1. ANCHOR-MH Weekly content list

| Week | Primary diagnostic category |
|---|---|
| 1. | PTSD |
| 2. | Borderline Personality Disorder |
| 3. | Bipolar Disorder |
| 4. | Autism Spectrum Disorder |
| 5. | Major Depressive Disorder |
| 6. | Chronic Pain |
| 7. | Perinatal Mental Health |
| 8. | Substance Use Disorders |
| 9. | Psychosis |
| 10. | FND |
| 11. | Physician Wellness |
| 12. | Course wrap-up |

**Supplementary Table 2. All responses to post-survey questions from Cohort 2.** Note that any identifying information has been removed but responses have not been edited for spelling or grammar.

| **Have you changed your approach to patient care in any way? (if yes, how?)** |
|---|
| - Yes, I feel much more comfortable discussing conditions chronic pain and borderline personality disorder from a physiologic perspective. I have many more resources to refer patients for e.g. ECT for treatment resistant depression and rTMS for PTSD.<br>- Yes, more likely to suggest particular psychotherapy for certain conditions. Have a better approach to medications for particular conditions.<br>- I think that I am better at explaining psychiatric conditions to patients in a less "hand waving" kind of way and feel more comfortable talking about the underlying biology.<br>- Yes, more empowered to discuss pathophysiology<br>- yes. I started looking for possible organic cause for my patient with psychiatric illness, more comfortable managing complex psychiatric presentations.<br>- Explaining mental health disorders using the neurobiological models/understanding<br>- This course has changed how I view the DSM and its limitations. I don't feel that I need to focus so much on ensuring my patients have the correct label, as their psychiatric illnesses rarely fit in those simple boxes. I am also more confident that what I am doing is correct but now I have a much better understanding of the pathophysiology of certain conditions and how to explain them to patients.<br>- I am way more aware that boundaries have to be set with patients in order to provide care. I liked [redacted] suggestion of using "and" instead of "but" for statements to avoid rejection<br>- Understanding the basis of some conditions that can be hard to treat leads me to be more understanding and empathetic towards my patients and I think will generally help reduce feelings of helplessness that can worsen burn out.<br>- Yes - more empathetic with “challenging” patients because of my greater understanding of neurobiologic mechanisms of psychiatric illness. More forgiving with myself when facing therapeutic challenges.<br>- Yes. I feel that I have a a more organized and empathetic approach to dealing with patients who have chronic pain, borderline personality disorder, and autism spectrum disorder.<br>- No - however I have better resources and understanding.<br>- Yes more patientce and empathy as understood neuroscience better and know more about community resources<br>- Yes. I understand pseudo seizures better. They are not seeking attention. There is an explanation of what is occurring with a treatment plan. I am more empathetic to this population, I am also aware of Access 24/7<br>- Yes, particularly for patients with borderline personality disorder. Found the BPD lecture very helpful.<br>- I am more capable of discussing trauma and it’s foundation for mental health concerns<br>- I try to explore a little bit more the possible underlying stressors leading up to their condition and offer more potential treatment avenues.<br>- Yes! I have a bank of resources I can turn to now for when I need answers to clinical questions. |

| |
|---|
| - One issue is I am more mindful of how my patient's emotions affect my own. That helps me reason more objectively about how I can help the patient. |
| **Do you have any specific feedback on the education sessions we ran in the first hour of each week?** |
| - I liked the small group format. I think the evaluation at the end has great learning value.<br>- Loved the videos<br>- Loved it.<br>- It would be useful to get the link sent out for the session in advance rather than by zoom as there are lots of circumstances where the zoom chat gets cleared<br>- It is well organized and common topics were adequately addressed.<br>- I really enjoyed the format of the first hour, however, there were a lot of great resources, more than I admitted have time to go back and review and read. The videos watched during the session itself were helpful.<br>- The sessions were excellent. I appreciated the videos, cases and many resources. Sometimes there was a bit of focus on urban resouces and programs, which is not applicable for rural physicians. I would also have appreciated some time to discuss the system issues and what we can do to advocate for change in our current mental health care system.<br>- I loved the videos. I thought it was an awesome session.<br>- Seeing sample cases and a general approach to management was very helpful. I wondered for more involved and common topics like PTSD , prenatal mental health and bipolar disorder if multiple cases and more time / more sessions would have helped me get even more comfortable with the management of these conditions. A heads up about the upcoming topic and resources sent out the night before to review.<br>- I really like the structure of it. Sometimes having 4 people in a group would be nice<br>- They were very effective. I appreciated the opportunity to discuss topics with peers. The resources were of high quality, especially the videos.<br>- I enjoyed the learning. Thank you so much for introducing wonderful resources. Having dedicated time to work (where we also get compensated for time) on our learning was so great!<br>- The sessions were truly mind-blowing and transformative. It deepened my understanding of the neuroscience underlying the most common mental health issues and highlighted their practical significance in clinical care. I also gained valuable awareness of available community resources and rapid access pathways, which will directly enhance patient management. The topics chosen were highly relevant and practical, offering immediate applicability in daily practice. Overall, this fellowship was an exceptional learning experience, and I am grateful for the opportunity to participate.<br>- I loved the discussion. The articles and videos were good. It was nice having different psychiatrist present.<br>- I really enjoyed these sessions!<br>- It was really great, I have learned lots and see even a greater need than ever before for patients to have access to trained therapist which we have so very little of in the current system.<br>- It was very informative and the format encouraged interaction, which was making the whole experience more rewarding<br>- None! They were excellent! |

| |
|---|
| - It might be wise to be clear about the core essense of how to diagnose and treat an illness - the essentials - vs. interesting recent research, emerging treatments, and speculation regarding pathophysiology. Both are interesting and helpful, but the first is essential and I think it is important to be clear what is being discussed. |
| **Do you have any specific feedback on the case conferencing component?** |
| - Awesome that so many psychiatrists are willing to take time to help us with these cases!<br>- I liked how the different psych specialties contributed. I appreciated the resource sharing. It was not easy to find cases weekly to share but thankfully there was always someone presenting.<br>- Given the emphasis on good quality therapy including trauma therapy etc, it would be interesting if there could be psychologists at the table too.<br>- I found this helpful<br>- to have further smaller groups and scheduled case presentation for the participants a week prior.<br>- This was also very useful and helpful to me and my patients I was able to discuss<br>- I really appreciated having the continuity of one psychiatrist with our rural group. I think it would be helpful to have a schedule for the family physicians for who was presenting which week as it did seem challenging to have a case ready every week. Also typically only 2 cases were discussed in the 1 hr session.<br>- Bigger groups to have a more robust discussion.<br>- having a set approach or expectations announced at the start would be helpful, ex. have participants enter preferred cases to discuss in the chat at the start of the session and people can decide which cases to cover 1st etc.<br>- I found it very helpful to discuss resources and next steps for patients (my own and others). My only complaint is that some of the resources I was referred to require a psychiatrist referral to access, which I didn’t know about until after contacting the programs<br>- I really enjoyed hearing perspectives and advice from the psychiatrists about our specific cases. I picked up a lot of 'clinical pearls' and felt that the psychiatrists were empathetic to the challenges we face as family physicians.<br>- I hope that group may get bigger as we get to the end of the session as people tends to run out of cases to discuss. I had one session where no one had any patients to discuss.<br>- The case presentations and discussions were the highlight of the program—engaging, insightful, and an epic learning experience. I greatly appreciated the contribution of all the psychiatrists involved, who were not only knowledgeable but also approachable and supportive. Their representation from various subspecialties of psychiatry provided a well-rounded and enriching perspective.<br>- I like doing the session in the summer. It would have been more difficult for me anyother time of year<br>- I discussed my concerns about the case conferencing directly with [redacted]<br>- It was very well done.<br>- The cases discussed were extremely difficult and the advice received from psychiatrist was very helpful<br>- The following points were brought up during my exit interview: - Possibly switching up the participants in the group every week to increase exposure to different types of patient populations. - Ensuring that the psychiatrists switch up every week to get exposure to different approaches to |

treatment. They each have a different niche / area of expertise and it would be nice to get exposure to all. Additionally, getting a heads up on which psychiatrists we have every week would be helpful from a case preparation standpoint.

- It might be good at the first session to suggest everyone who has a case start the session by submiting a short summary in the chat.

**Semi-structured interview guide**

All interviews were conducted via zoom. Interview transcripts were recorded using otter.ai. Individuals were assured of anonymity and confidentiality, and they were told they could contact the interviewer afterward if they had any concerns about what had been said, or if there was anything else they wanted to add or remove (no additional contact occurred). Afterwards, transcripts were cleaned and any potentially identifying details were removed as necessary (requests, e.g., for additional contact were maintained).

Interview questions:

1. What were the strengths of this experience for you, and did you meet your goals?

2. Did you experience any epiphanies or "lightbulb moments"?

3. Is there any way in which this experience has changed your practice or improved patient care? If so, how? (e.g., increased comfort? increased skill?) Any outcomes you can point to?

4. Do you find you think differently about psychiatric patients now? Or have you been able to help some of them think differently about themselves (stigma)

5. Is there any ongoing follow-up you'd like to see?

6. If we were to offer this experience again, do you have any suggestions for how to improve it?

7. Anything else you'd like to add?

**Cohort 1 thematic analysis additional quotes relating to themes**
***a. Increased confidence***
Most participants agreed the course content made them feel more confident in tackling the kinds of psychiatric issues they are likely to encounter everyday.

- One thing, I feel more confident and less scared.
- I think more confidence in saying, like, “Okay, I think we're on the right track. I don't think there's anything else that we're missing.” So that's that's been useful… I think definitely a difference in clinical decision making, reduced referrals, and then supporting my colleagues. I think those are the three big things.
- Yeah, I think my diagnostic clarification for sure. I don't even know if it improved, but I definitely have more confidence in it, in making diagnoses, and then also in treating.
- I feel a bit more empowered and just kind of up-to-date in my approach.
- I think what they would try and do is teach you about each condition in such a way [as to] allow you to explain it with patients.
- I've come to understand that it goes way beyond prescriptive prescription or prescribing some of the psychotropic medications. [For example], the role of lifestyle during the course… it was an eye opener, because I realized, “oh, you know, I should take my lifestyle and dietary counseling more seriously,” and even just try to link it during my consultation with the patients and say, “hey, there's a link between this and this.”
- I think, just knowing, like, what the resources and guidelines are, so when I do have [a specific disorder in front of me, knowing] what are the first line and second line [treatments], and having that quick resource to go to.
- [There] was also the fact that I became more comfortable with adjusting doses [beyond the generally accepted limit, in certain cases].
- I think that allowing for access of resources… [building] confidence when managing different psychiatric problems was helpful.
- It was really, was very impactful. This was the best course so far, like, I've taken. It was very intentional, well prepared, well thought through. So, I think I’d highly recommend this to any of my colleagues.

***b. Making like-minded connections and understanding the mental health landscape***
A related positive finding was that they felt they had a better sense of how to proceed and whom to contact when complex cases crossed their desk. Many commented on how gracious and helpful the team was, and how supported they felt in getting information.

- Access to the psychiatrist and being able to discuss difficult cases directly with the specialist; knowing the fact that we were able to talk to them once in a week, was a big, big, plus.
- I think these connections are so vital, right? Like, you kind of practice in your silo, and you kind of forget, “oh, everybody else is going through the exact same thing or struggling with the same things.”
- The major strength of the program was the interactions, right, like talking about different issues and getting an approach right, and I'm able to communicate with specialists. That

was wonderful, right? And it was really good to kind of get some information about the resources in the locality.

- The one thing I was just so amazed at, like, just brainstorming, yeah, I was so amazed at all of our colleagues, like it was so generous, like the psychiatry colleagues being there and, like, you know, sharing their knowledge and everything. It was just so valuable.
- Again, kind of building on those relationships that we just started to establish - I think that's really valuable in the healthcare system. To know your local psychiatrist, to know the local services, and to share information about their perspective, our perspective. Those are some of the intangibles that come out of this stuff. And so that would motivate me to keep going with it.
- All of the psychiatrists that were engaged with the program were excellent. They had great, great answers to everything, great perspectives. They were very, like, collegial. I loved all of them.
- So, it's nice to kind of surround yourself with like-minded [people]… that had that same passion, and then the resources are really greatly received between everybody. And everyone was really good, like, if they found additional topics or things, they would share them. It wasn't like this, competitive [atmosphere]. It was more like just a sharing of knowledge, which I really liked.
- So, we got to know some of the psychiatrists and some of the other physicians who are quite like-minded in terms of supporting clients who have mental health challenges. I thought just the fact that it was a cross departments… initiative, I thought that was really great.
- I liked the amount of facilitators and psychiatrists that we had access to. I think if we had such a large group of Family Physicians and, like, one or two psychiatrists, it might have felt overwhelming or inappropriate in terms of level of support, but I thought that it was really well staffed.
- What I loved about it was the opportunity to connect with, like, colleagues and psychiatrists.
- It allowed me to have a better understanding of the landscape of Psychiatry in Edmonton.
- Some of the refractory cases, [as] I called them, I was really able to find a path how to help them and to allocate the resources and to direct them to the to the specialists.
- It was very collaborative and I think in the end, it will impact patient care in a really positive way.

***c. Impressive educational resources***
Participants were generally very impressed by the breadth and depth of the educational resources, although some felt they might have been a bit too heavy or high-level. However, they appreciated having defined takeaways ("pearls") that provided helpful, memorable direction.

- So, I loved learning about some of the like actual neuroscience behind some of these psych disorders. I don't think we learn a lot about it in med school or residency, and so that's going to be helpful when trying to explain to patients.

- I liked the breadth of case presentations and teaching that we did, so I felt like we covered a lot of important topics that I had questions on… in terms of, like, videos and, you know, access to the literature and things like that - I thought it worked really well.
- No, it was amazing. It was amazing. And everybody was so helpful and kind to guide us regarding the cases, as well as give us directions as well as resources available that can be used.
- Loved them. So great.
- The background reading that was given to us helped to boost my confidence in some of the choices I've been making already. And then new things that I learned along the way, right? So in that sense, I would say that that was a strong point of it… The vignettes that we had to review, like the video, even some of the cases that they used as examples for us to answer - they were very practical. So, I appreciated that.
- What the educational session brought in tons of information and pearls of wisdom from specialists that we really spoke to, and it was really good. And the best thing is also that we can take it to the discussion after that. And, you know, the resources, the information, the guidelines provided were really great.
- I felt I picked up a lot of information, even if I wasn't, you know, reading kind of pages of things, it's "the pearls" formatting was really helpful.
- It's robust the quality of the literature that was given us. The resources were great.
- I also like that it worked us through on, like, a timeline. So, like, we didn't go down rabbit holes … I love that there were different, like I said, multi modal approaches. So there was videos, there was papers that we could scan, there were the case studies that we would work through. And, yeah, so I thought, I thought they were excellent, and it was very well facilitated.
- You're jamming a lot of content into an hour. You don't have a ton of time to really digest it.
- I think just that I'd be curious to know how they chose the subjects that they did each week. If it was stuff that was noted from psychiatry being like, "we feel like these are gaps," or if it was feedback from Family Physicians saying, "these are our gaps"…
- I think only one or two sessions, the didactic sessions, was there maybe a little bit more, like scientific articles with research. And I was like, "oh, it's not as quite as practical." Because, like, it wasn't practice-changing.
- Sometimes the cases in the education sessions would be like, "they've tried the first four options, and I'm having to remind myself what the first four options are…" Like, it'd almost be nice to have almost like two sessions, like one for each level, like one just for, like, here's the review of the basic stuff, right? And then a second session would be needed for, like, when that basics isn't working.

***d. Case conference section somewhat uneven***

Predictably, individuals who had specific cases to bring forward found the greatest value with the case conferences where they could ask specific questions and get timely feedback. A few respondents expressed "fear of missing out", wondering what was transpiring in other groups…

- I think like just being armed with some more knowledge [has been helpful], in, you know, for specific cases and whatnot.
- I did, like present two or three different scenarios that I was going through in the practice to kind of get a sense and the feedback and the information that gave, right? I use that to further manage the patient, right? So yeah, it has been really a good experience, fantastic.
- I found the case conferencing in our group… with the psychiatrist right there, that was so helpful. Because it was family doctors bringing cases that we see and having to being able to ask very, like, specific questions, or even, you know, just targeted questions to kind of get us to the next level.
- That's one of the strongest suits, because I got to discuss three cases I remember that were keeping me up at night.
- I think having more family docs in that group would have been also nicer to bring [a greater] variety of cases.
- I work in a very kind of sub specialized area… like, four or five standard deviations away from what a lot of other practitioners see in general practice. And so sometimes I found that [for] some of the really tough cases I'm grappling with, I didn't feel like there was enough support. [Similarly], I kind of started to feel like the other clinicians in the room felt like it was a bit too far removed from their own experience.
- Um, I think there's a little bit of FOMO where - because it's very specific to the cases in that hour - you were kind of wondering what different experts were in different rooms, with different cases, some stuff there…
- I understand the rationale of keeping the groups the same every week, but I definitely had some, like, FOMO about, like, what other groups were talking about, so I don't know. And it's hard, I'm sure, with the first time to know if it would be better or worse, but I would definitely not be opposed to switching up the groups week to week.
- I had just started at this new clinic, and I had less complex psychiatric [patients], so I didn't actually have very many conflicts [and] I wasn't able to ask as much personal questions related to a patient case, which was unfortunate.

**Cohort 2 thematic analysis additional quotes relating to themes**

***a. More confidence seeing psychiatric patients***

- Really, I have more resources and bit more confidence going ahead with the treatment strategies.
- All the psychiatrists were really helpful. And yeah, in the sense that, you know, I think I'm more confident, you know, treating mental health issues. I met my goals, so yeah.
- So yeah, like I think, just overall learning more about the treatment modalities, and just being more connected with the resources has been a lot more helpful I guess in improving my confidence in my approach to these patients.
- [My goal] was just mostly to feel more confident in managing more complex cases that are kind of beyond first line measures. And I do definitely feel more equipped for some of those more, like, just more complex cases.
- SSRIs and stimulants, those feel like easy to start medications. But even like atypical antipsychotics for aggression, or, like lithium for bipolar - those kind of things [seemed] a little bit more unapproachable. And I definitely feel like I had the learning to make those feel less scary, to prescribe. SSRIs and stimulants, those feel like easy to start medications. But even like atypical antipsychotics for aggression, or, like lithium for bipolar - those kind of things [seemed] a little bit more unapproachable. And I definitely feel like I had the learning to make those feel less scary, to prescribe.
- I think also just seeing that it's such an art form. Like, you would ask one psychiatrist one thing and the other, and they would both have different approaches. That there is not one right way, you know, I think that was really nice to learn. Because sometimes I think you're like - you know, you call a consultant, [and they're] like, "Well, I would have done this." And literally, to see two psychiatrists talk in the space, like, "oh, I would do that" and "I do this," it really gave me a bit more confidence to be like, there isn't one [correct way]. You know, you just have to pick an attempt and get on board with your patient, and then, yeah, if it doesn't work, you try again. And I feel more confident now knowing that they do that too.
- There were some affirmations as well. So, it was great to hear from the psychiatrist. They were like, "Oh, you're managing these really complex patients out in the community." And so it was great to get that validation for, you know, honestly, doing more than people expect us to be doing, or realizing that we're doing.
- Now it's more efficient to actually see the patients, because you know what you're seeing. You now can express what you're seeing. You can then explain that to the patient, what the way. And now you have a referral place to send them to. And there's a lot less reinventing the wheel for these patients. You now have your little algorithm on where to go, and that makes -, internal anxious goes down.

***b. For many, the program has changed practice***

- [With] someone who has been getting treatment, [but] not getting better. what I was thinking [before] was more like maybe trying different medication, or increasing the dose, or maybe getting them referred to a psychiatrist. But then I realized now I might need to do more  workup to see - get into their metabolic panels, and see if they have maybe some sort of like a hypercorticism, or [are] in a prediabetic state, or I even started one patient after finding out that the patient has metabolic abnormalities, and started treating them in that way as well.

- One of the weeks that stood out is when, when we had a group, sort of study on chronic pain, and that's changed my approach and how I talk to patients about their pain and their goals. You know, it's we often get into this sort of cycle, or this rut where patients are hoping that there's a test or treatment there that's going to just make it all better… [I learned] it's helpful to explain to them that, you know, if your pain is on a scale from one to 10, and that's always a nine, that we need a whole bunch of things cumulatively together to each bring it down by one point. [Gives helpful detailed explanation.]
- So, I guess I wasn't as much pushing towards Cognitive Processing Therapy. Obviously, I knew that they exist, but like the first line evidence was SSRI, so [I'd] always start on SSRI and then do CPT at the same time. And then the, I think, PCL-5 questionnaire too, that's also helpful in kind of assessing, right? So, I've never done that, given that questionnaire to my patients before, but I started doing that. And then also I encouraged patients more about the CPT and SSRI in kind of trauma therapy. So, I guess that's one of the more pivotal [changes] in my practice.
- That was great, you know, to kind of have those discussions and get the feedback from the psychiatrist involved with the case conferences part, and I kind of relay[ed] that to my patients, and they were actually thrilled, and they were actually happy to do the changes. And we'll see how it goes.
- One example was one patient with just got a lot of situational stresses in terms of husband passing away, recently retired, diagnosis of breast cancer, now with, like, really bad hip pain - just like a whole bunch of stuff came all at once for her. And she's got alcohol use disorder, she went through this like hip replacement, and ended up with a lot of pain while waiting for the hip replacement, and so we needed to use opioids to kind of manage that pain in the interim, with the idea and knowledge that this might be something that's going to be tricky coming off of surgery. So, managing her overall pain, overall mental health, addictions, through the support with some of the peers and psychiatrists like we were able to kind of get her off her opioids. And she was very, very motivated. And it could have been a completely different situation, but at least having like, some basic understanding of, like, what to do and what [we] tried to not do was super helpful and very timely.

***c. One key learning was that treatments were more readily accessible than they would have thought*** (this was considered analogous to the "Understanding the mental healthcare landscape and making connections" theme identified in cohort 1 by three raters who were not involved in the initial generation of themes)

- Yeah, I didn't, like, ever I don't think, refer patients for ECT or rTMS. And now I'm like, "Oh no, these are actually really great treatment options for those treatment-resistant patients."

- So being introduced to all those resources, I've already utilized some of the resources, like sending, you know, young adults to Kickstart, for example, as like a resource. Knowing that I can refer patients for, you know, rTMS.
- I think one of the [lightbulb moments] was like, not being afraid to send patients for ECT. That was something that I always thought was the modality that was kind of like outside of what's accessible for family doctors.
- I've already had one patient that I would never in a million years have thought to send for outpatient ECT - I've already had one I've talked to, I would have never done that. (*Inquired whether that went well.*) Very, actually. So, we are kind of pulling back some of her meds, because she's, like, total polypharmacy. But I planted that seed that, "you know, what? If we can't get you better, this would be a great option for you." And she's, like, would be perfect for it. It was great. I have no complaints.

***d. The neuroscience resources were very effective and valuable to have at hand*** (this was considered analogous to the "Well-received educational resources" theme identified in cohort 1 by three raters who were not involved in the initial generation of themes)

- The other biggest, I think, strength of the program was being provided with so many really reliable psychiatry vetted resources that I can access because I think that's part of the challenge of being a family doctor is obviously you're not going to know everything, but knowing where to look is really huge. And so just being given, like this whole collection of resources about a lot of different issues, I found to be very valuable.
- And the best thing is the resources that they talked about right, [a] lot of them that I was not aware of. So, I feel so hopeful now that I can assist my patients in a better way.
- I really liked how this fellowship tied neuroscience aspect to make sense of the mental health condition, because I think that was quite lacking during my training… So it was actually quite refreshing to see the scientific evidence behind it, like, how far we have come along.
- Like all that neurophysiology that we can turn around and say to patients, "you know, what: This, this, this." Whereas before, you know, you didn't really, necessarily have that background. You would just give them basically a DSM [description], right? Yeah. So, it was nice to have that neurophysiology in the background. I think the videos are what stuck with me the most, to be honest, and how they tied into the cases, which was wonderful.
- I really liked the incorporation of a lot of the papers that they provided, because I think - especially as teaching [tool] - that was stuff that I could give to students and, like, give them in a way to, you know, challenge [stigma] without just being like, "you should think differently about this." So, I found the resources super helpful.

***e. "Clinical pearls" and a-ha moments were numerous*** (this was considered analogous to the "Well-received educational resources" theme identified in cohort 1 by three raters who were not involved in the initial generation of themes)

- Yeah, I think lots. Probably one [per] session, I would say.

- I mean, in every session. In every session, and especially the last one - when they're talking about the physician stress and all those kind of things are burnout, that is like… [an] epic session. And I was feeling like, they are so much in touch with the reality, the whole program, of what family physician are going through. And I think it is an excellent, excellent program for most of the family physicians, and I [will] keep on promoting to my colleagues.
- Yeah, I think there, there were some of those every single week, kind of clinical pearls that I hadn't really thought about or been aware of. One example is like in a recent session, we were talking about adult ADHD and how it can be challenging to, you know, feel really confident about that diagnosis and how to proceed with prescribing. And so it was, it was mentioned that, hey, it's really useful if you can try to get some collateral history from someone that knew this person when they were a child, and get that documented,
- Just the effect of epigenetics and like how that kind of plays into a lot of these different psychiatric conditions was - I felt like that was kind of eye opening.
- Well, there were a bunch of things that I did not know. So, for example, like the genetic basis and genetic testing for autism, like that was a brand new one for me, right? Even the opioid use disorder module, they talk about the brain changes, etc, etc. I did not know about that, so there were lots of aha moments.

***f. Helping patients see mental illness as medical can be important to their recovery***

- I think that might be helpful for patients to like, sort of, you know, not view their condition as like a personal failing, but as like a medical condition, just like any other medical condition.
- It's like you not having, you having all of these traumatic things happen to you, and it like actually rewiring your brain. And I think that thinking of psychiatric illnesses in that way is kind of helpful. It feels like a more treatable entity to me, and I think for patients, it makes them feel less guilty about having some of these diagnoses. It's not like a "Oh, it's not purely me just not being able to cope or not having XYZ. It's actually like some changes that are happening to my brain and, like, obviously you can't control that."
- Well, I think that it was nice to see that all of the commonly-seen, like, issues in mental health - and even including, like, borderline personality disorder, including chronic pain - that there are differences and neural pathways that we can see. Like that the chronic levels of stress and chronic or repetitive trauma has, like, a real impact on how the brain changes and processes things. So, it was really nice to see that, and to be able to tell patients, "no, actually, yeah, like, your brain has been wired differently now. And like, we just… what we need to do is help kind of reset everything, and, like, rewire things so that you process things in a in a more manageable fashion," right?
- I have a patient that I've known for a long time in our practice, shortly after one of our sessions that we reviewed about PTSD, I think I was able to kind of explain a bit more how the signals in her brain change and how you know why the certain type of therapy would be more beneficial for her. And I think it was a moment where, like, I would never

have gone into this detail of a patient before. And I think she really appreciated it, and I think it motivated her to actually pursue, you know, the proper PTSD counseling and therapy that she would normally maybe not have done.

- Yeah, I think that the, like, the traditionally, "it's all in your head" to sort of things like, yes, while it's technically literally in your head, you know, it's not just something that people are just making up. And being able to say that, like, with just solid confidence is really nice, right?
- That [newer research] helps a little bit when you're trying to talk with a patient suffering from these conditions, or like, "why is this happening?" "Well, here's some… our current understanding of these processes." It doesn't necessarily lead to new treatment modalities, but at least you can validate people's experience a little bit, if you can give a bit of info on why, why they're feeling what they're feeling… that, I think, helps people understand what's going on a little bit. Yeah, it's frustrating. It [chronic pain] can be very challenging to treat, but I think being able to kind of point to some of the physiology behind these conditions does help a bit.
- I think it's and maybe it's just changed kind of my own internal understanding of things, and so I think the way I speak to my patients is a bit different now, and hopefully in a positive way, where just kind of trying to build as my understanding has deepened of mental health that I'm trying to impart that on them. I think that's probably the main thing, and is that that's changed.

***g. Many noted there was an actual shift in their language when speaking to patients***

- I think when you can have a language to explain to patients, you know… it changes how their brain processes the information, and how they're able to cope with it… I think having that also empowers patients to better understand that it's not just, you know, "there's something wrong with me" or they're a bad person… So just having, just hearing some of the language that's used, and then being able to apply that in a way that patients can understand, I think, is also very helpful.
- I think there was a lot of interesting ways of phrasing and presenting some of the information, for example, around functional neurological disorders come to mind where we're presenting things in slightly different light…
- Even some of the language, working through some of the cases, on how to describe, you know, the neurobiology of different conditions and explain to patients and have the language to use was super helpful, and I definitely applied that pretty quickly.
- Because of the course, they gave me a vocabulary to actually speak to the patients with. I don't know if it's proper terminology, but more – a better explanation of what they're going through, right? Which can relate to that a bit more. I think the conversation is more refined, right?
- So I think having that language, and I really liked the language around, like the mental health jar, in terms of explaining, like your genetic predisposition, but also kind of like the life stressors and situations and then your coping mechanisms, and just having those analogies, I think another one was like blueberry muffin recipe or something like that, to

just explain, like, you know, “these are the tools that we have, and you know, this is what we can help us.” So just having, just hearing some of the language that's used, and then being able to apply that in a way that patients can understand, I think, is also very helpful.

***h. Medical doctors themselves came to understand their own stigma, increasing their patience***

- It was like [a] really mind-blowing session when we understand that it's not under their control. It's not just seeking attention. It’s something [neurological] going on. So, those kind of things were really [eye-opening] and it will definitely help my patients and my perspective as well, and probably helping their families as well, to understand. Because we all are human beings and [we] start losing patients when we don't see any success or any improvement.
- I think that, like, just learning a lot more about some of the things that are… that I would find more challenging to treat. I feel like I'm a bit more - like, I have a bit more patience with it. Like, on my own end, you know, I'm not as frustrated. Like, “okay, what do I do now?” Like, this patient isn't getting better, but maybe now I'm… I can kind of know that there's other options out there, without necessarily having to directly refer and kind of wait.
- I really liked when they were able to explain, like, kind of the science behind it, too, with those little mouse models. That really helped me. Because sometimes, like with Borderline patients, I would just be like, they're kind of annoying, right? Like they've kind of chosen that for themselves. They've chosen that path… But they didn't really choose that path.
- I think, I think the experience of the last few weeks has probably helped me be a little bit less judgmental or jump to conclusions about, you know, what patients are thinking and feeling. And I'll make more of a conscious effort to try to validate, you know, a lot of their concerns that are, I guess, you know, subjective in a way where there's not a physical test that that can prove or disprove what they're saying… there's actual, you know, changes in the brains of these people that that is very real, and I haven't really considered that before. So, yeah, I think it's, it's helping me be a bit more open minded and less judgmental.
- Sometimes you - we do get frustrated with patients, and we're like, “Come on. Like, did you really need to go to the ER fifteen times for this,” right? But when you add a little background to it, and start to think a little bit more of, you know, child abuse, and their childhood, and things that we don't always bring to the forefront when we're running a busy clinic, right? And we're like, “we’ve got to move on,” right? Just, it was good to hear, was good to see some of that stuff and just… a little bit more compassion.
- Also, you know, I've always thought about borderline personality disorders, as yet another illness where, you know, I kind of moan and groan… I almost don’t even want to tell patients they have borderline personality disorder, because *I* have such a negative stigma of borderline personality disorder, I forget the patients sometimes just want to have a diagnosis… So it was good to again have that conversation, to say, “Hey, you know, for some patients, you know, the stigmas we carry on in our brains are not

necessarily what the patient experiences, and withholding that label actually causes harm."

- I think now just having that better understanding and more empathy… it just reinforced that, like, this is that, yes, they are difficult to manage, but at the same time, like, there's more reasons of when you understand the biology of it, then it just gives that more compassion and empathy.

***i. Beyond increasing patience, some pointed out that having that understanding actually decreased burnout***

- Well, the thing is, when you see somebody, and you really – and you know that something's wrong, but you don't know what to do with them - then you're busy thinking of what do you do with that? And that gives you a lot more stress. You're trying to read around it, trying to figure out who to refer to, and you're trying to help your staff calm down. These people rile up her staff members. All that takes much more time.
- You kind of develop a feeling towards [these patients] and you feel helpless and you feel frustrated, but looking at it from a perspective of, "Where is the biology? Where is the physiology of it? What might be some of the reasons why someone would present this way?" And this was a similar theme in with cases like personality disorders or PTSD and like, "what are the driving forces of what gets people there," which I don't think has been previously really well taught in medical school... And part of it is it's just not really super well understood, but to have, like, some kind of physiological basis of some of these things, it's like it's, I think, almost protective in ways of like, preventing burnout in physicians being able to say, "hey, it's not necessarily me. It's this sort of pattern that can be seen in these conditions, and so then, similar to other patterns that we see, these are the algorithms, or these are the treatment pathways then we can take."
- Sometimes it can be tricky to recognize, but kind of knowing and acknowledging that it's there and knowing that perhaps there's like a biochemical basis to some of these presentations, and there's a pattern of behavior helps you not only recognize, but sort of understand and empathize a little bit more, is that some of these people aren't just being difficult, you know? And just like I say to my toddler, like they're they're not giving me a hard time, they're having a hard time. And like, understanding the neurophysiology, or, like, at least knowing that there's some sort of likely connection or basis makes it like so that you're a little bit more forgiving and maybe a little bit more understanding and empathetic, and so with that like you don't feel as run down.
